# Diagnosing Cognitive Disorders in Older Adults with Temporal Lobe Epilepsy

**DOI:** 10.1101/2020.09.05.20189084

**Authors:** Anny Reyes, Erik Kaestner, Emily C. Edmonds, Anna Christina Macari, Zhong Irene Wang, Daniel L. Drane, Vineet Punia, Robyn M. Busch, Bruce P. Hermann, Carrie R. McDonald, for the Alzheimer’s Disease Neuroimaging Initiative

**Affiliations:** Center for Multimodal Imaging and Genetics, University of California, San Diego, CA, USA; San Diego State University/University of California San Diego Joint Doctoral Program in Clinical Psychology, San Diego, CA, USA; Department of Psychiatry, University of California, San Diego, CA, USA; Veterans Affairs San Diego Healthcare System, San Diego, CA, USA; Epilepsy Center, Neurological Institute, Cleveland Clinic; Departments of Neurology and Pediatrics, Emory University School of Medicine, Atlanta, GA, USA; Department of Neurology, University of Washington, Seattle, WA, USA; Department of Neurology, Cleveland Clinic; Department of Neurology, University of Wisconsin School of Medicine and Public Health

**Author notes:** Corresponding Author: Carrie R. McDonald Ph.D. Altman CTRI building; Floor #: 4W 9452 Medical Center Drive, La Jolla, CA 92037. Data used in preparation of this article were obtained from the Alzheimer’s Disease Neuroimaging Initiative (ADNI) database (https://adni.loni.usc.edu). As such, the investigators within the ADNI contributed to the design and implementation of ADNI and/or provided data but did not participate in analysis or writing of this report. A complete listing of ADNI investigators can be found at: http://adni.loni.usc.edu/wp-content/uploads/how_to_apply/ADNI_Acknowledgement_List.pdf.

## Abstract

**Objective:** To characterize the nature and prevalence of cognitive disorders in older adults with temporal lobe epilepsy (TLE) and compare their cognitive profiles to non-epileptic patients with mild cognitive impairment (i.e., *classic* MCI; cMCI).

**Methods:** Seventy-eight older patients with TLE, 77 cMCI, and 69 normal aging controls (NAC), all 55-80 years, completed neuropsychological measures of memory, language, executive function and processing speed. An actuarial neuropsychological method designed to diagnose MCI was applied to individual patients to identify older adults with TLE who met diagnostic criteria for MCI (TLE-MCI). A linear classifier was performed to evaluate how well the diagnostic criteria differentiated patients with TLE-MCI from cMCI. In TLE, the contribution of epilepsy-related and vascular risk factors to cognitive impairment was evaluated using multiple regression.

**Results:** Forty-three TLE patients (60%) met criteria for TLE-MCI, demonstrating marked deficits in both memory and language. A classification model between TLE-MCI and cMCI correctly classified 81.1% (90.6% specificity, 61.3% sensitivity) of the cohort based on neuropsychological scores. Whereas TLE-MCI showed greater deficits in language relative to cMCI, patients with cMCI showed greater rapid forgetting on memory measures. Both epilepsy-related risk factors and the presence of leukoaraiosis on MRI contributed to impairment profiles in TLE-MCI.

**Interpretation:** Approximately 60% of older adults with TLE meet diagnostic criteria for a cognitive disorder associated with aging (i.e., MCI). The TLE-MCI phenotype may be secondary to an accumulation of epilepsy and vascular risk factors, signal the onset of a neurodegenerative disease, or represent a combination of both factors.

## Introduction

Older adults represent the fastest growing population of patients with epilepsy^1^. These patients present with a multitude of risk factors for accelerated cognitive and brain aging, including vascular/metabolic risk factors, altered lifestyles, and poor quality of life^2–4^. With a globally aging population, it is critical to fully characterize the cognitive impairments present in older adults with epilepsy and to identity those at increased risk for accelerated aging and progression to dementia.

Temporal lobe epilepsy (TLE) represents the most common form of focal epilepsy in adults^5^. Despite numerous studies demonstrating pervasive cognitive deficits in young-to-middle aged patients with TLE^6, 7^, the cognitive profiles of older adults has not been comprehensively characterized^2^. Furthermore, the prevalence of cognitive disorders in older patients with TLE is unknown. In a subset of epilepsy patients *at risk* for accelerated aging, cognitive impairments have been described as similar to those in classic mild cognitive impairment (cMCI)^8, 9^, the preclinical phase of Alzheimer’s disease (AD), including prominent deficits in memory. However, these studies included small samples, a wide range of epilepsy syndromes, or only compared the average cognitive profiles across groups.

We provide the first systematic characterization of the nature and prevalence of a cognitive disorder in a large group of older patients with TLE and directly compare their cognitive profiles to those with cMCI and normally aging controls. Given the high prevalence of cerebrovascular risk factors (CVRFs) in epilepsy and their impact on the aging brain, we also evaluate the contribution of both epilepsy-related clinical factors and CVRFs to cognitive impairment. We hypothesize that a sizable proportion of patients with TLE will meet criteria for a cognitive disorder and that their performance will be comparable to patients with cMCI. Based on the existing literature, we predict that both clinical factors and CVRFs will contribute to the extent of cognitive impairment observed in patients who meet criteria for a cognitive disorder.

## Methods

### TLE Patients

This study was approved by Institutional Review Boards at University of California, San Diego (UCSD), Emory University, Cleveland Clinic, and University of Wisconsin-Madison (UWM). Written informed consent was obtained from all TLE patients at UCSD, Emory, and UWM. At Cleveland Clinic, data were collected as part of an IRB-approved data registry. Seventy-eight older patients with TLE met the following inclusion criteria for the study: between 55 and 80 years of age, treated at a Level 4 epilepsy center, and diagnosed with TLE by a board-certified neurologist with expertise in epileptology, in accordance with International League Against Epilepsy criteria^10^, using video-EEG telemetry, seizure semiology, and neuroimaging evaluation. No patients with TLE had a known diagnosis of MCI or dementia at the time of their neuropsychological evaluation.

### cMCI and NAC groups

Data for the cMCI and normally aging controls (NAC) groups used in the preparation of this article were obtained from the Alzheimer’s Disease Neuroimaging Initiative (ADNI) database (https://adni.loni.usc.edu). The ADNI was launched in 2003 as a public-private partnership, led by Principal Investigator Michael W. Weiner, MD. The primary goal of ADNI has been to test whether serial magnetic resonance imaging (MRI), positron emission tomography (PET), other biological markers, and clinical and neuropsychological assessment can be combined to measure the progression of MCI and early AD. For up-to-date information, see https://www.adni-info.org.

Seventy-seven patients with cMCI and 69 NAC between the ages of 55 and 80 were selected from the ADNI database to match the TLE sample as closely as possible in age and sex. Criteria for ADNI eligibility and diagnostic classifications are described at http://www.adniinfo.org/Scientists/ADNIGrant/ProtocolSummary.aspx. All participants were determined to be non-demented by ADNI, and dementia was determined based on NINCDS/ADRDA criteria for probably Alzheimer’s disease. None of the patients with cMCI had a history of other known neurological disorder, including epilepsy. Following recruitment and ADNI diagnosis, all participants completed a battery of neuropsychological tests at baseline and yearly thereafter.

### Neuropsychological measures and clinical variables

Measures of episodic memory included the delayed recall and recognition trials from the Rey Auditory Verbal Learning Test (RAVLT-Delayed and RAVLT-Recognition, respectively). Measures of language included visual confrontation naming with the Boston Naming Test (BNT)^11^ and semantic fluency with the Animal Fluency (AF). Measures of graphomotor speed and executive functioning included Trail-Making Test Parts A (TMT-A) and B (TMT-B), respectively. Language and graphomotor speed/executive function scores were corrected for age, education, sex and race based on normative data from the expanded Halstead-Reitan Battery^12^. Scores from the RAVLT were age corrected using the Mayo Older Americans Normative Study^13^. Of note, the Mayo older adult norms do not include total AVLT learning and therefore, we only included delayed recall (RAVLT-Delayed) and recognition (RAVLT-Recog) in our analysis. The same norms were used for all three groups (cMCI, TLE, and NAC). For TLE only, LM1 (immediate recall) and LM2 (delayed recall) from the WMS-Third Edition (WMS-III)^14^ were also included for diagnostic purposes. LM scores were corrected for age using the norms provided by the test manual. All scores were converted to T-scores (mean = 50, standard deviation = 10) for consistency and ease of interpretability.

For all TLE patients, epilepsy-related clinical variables and CVRFs were collected during standard clinical examination. CVRFs included diagnosis of hypertension, hyperlipidemia, diabetes mellitus, and/or obesity defined by a body mass index (BMI; mass [kg]/ height [m]^2^) ≥ 30. The presence of leukoaraiosis and mesial temporal sclerosis (MTS) were determined by inspection of MRI images by a board-certified neuroradiologist for clinical purposes. Leukoaraiosis was determined based on visual analysis of T2/FLAIR hyperintense foci in the periventricular/subcortical white matter. For the purpose of this study, MTS and leukoaraiosis were included as binary variables.

### Neuropsychological diagnostic classification

There is currently no consensus on a diagnostic definition for cognitive disorders in epilepsy, especially as they intersect with the most common disorders of aging. In an attempt to arrive at diagnostic criteria that can be used in older adults with TLE, we applied a comprehensive neuropsychological method (i.e., Jak/Bondi diagnostic classification) that has been recently used to improve diagnosis and classification of MCI cohorts^15–17^. These criteria are based on objective measures of cognition and offer an operational definition of impairment. The following tests were used for TLE patients: BNT, AF, TMT-A, TMT-B, LM1, and LM2. Patients were considered to meet criteria for *TLE-MCI* if any one of the following two criteria were met: 1) they had impaired scores, defined as > 1 standard deviation (SD) below the demographically-corrected normative mean, on two measures within at least one cognitive domain (i.e., memory, language, or speed/executive function); or 2) they had one impaired score, defined as > 1 SD below the demographically-corrected normative mean, in each of the three cognitive domains sampled. Seventy-one patients (91%) had sufficient cognitive data and were therefore included in all subsequent analysis.

To ensure comparability, ADNI cMCI and NAC participants were also classified using the Jack/Bondi criteria^16, 17^ (described above). Diagnostic classification was based on the following tests: BNT, AF, TMT-A, TMT-B, RAVLT-Delayed, and RAVLT-Recog. None of these tests were used to determine initial ADNI diagnostic classification. Sixty-four (83.1%) participants classified as cMCI based on the ADNI criteria met criteria for cMCI based on the Jak/Bondi criteria. The other 16.9% of patients were classified as cognitively normal, likely representing the *false positive* cases identified in previous ADNI studies^17, 18^. Sixty-five (94.2%) NAC participants classified as cognitively normal based on the ADNI criteria were classified as cognitively intact based on the Jak/Bondi actuarial neuropsychological criteria. The remaining participants, as well as the false positive cases, were excluded from subsequent analyses.

### Statistical analysis

Analysis of variance (ANOVA), independent *t*-tests and Fisher’s Exact tests were used to test for differences in clinical and demographic variables and neuropsychological performance when appropriate. Analysis of covariance (ANCOVA), controlling for age, sex, and education were conducted to compare neuropsychological performance (T-scores) across groups. When results from the ANCOVA were significant, group contrasts were assessed using post-hoc pairwise tests with Bonferroni correction. Multiple comparisons were corrected using Benjamini-Hochberg false discovery rate. Stepwise linear regressions were conducted to evaluate the contribution of demographic, epilepsy-related clinical variables, and CVRFs to cognitive performance. Finally, a discriminant function analysis (DFA) was performed to test whether neuropsychological profiles could correctly classify TLE-MCI and cMCI at the individual subject level.

### Data Availability Statement

Authors have full access to all study data and participant consent forms and take full responsibility for the data, the conduct of the research, the analysis and interpretation of the data, and the right to publish all data.

## Results

### Demographic characteristics

Table 1 includes demographic variables across groups. There were differences in age across groups; NACs were older than patients with TLE (*p* < .001) and cMCI (*p* < .001), and patients with cMCI were older than those with TLE (*p* <.001). There were also differences in education across groups; cMCI (*p* < .001) and NAC (*p* < .001) had more years of education relative to TLE. There were no differences in sex across the groups. Therefore, age and education were included as covariates. Given the expected effects of sex on some neuropsychological tests (e.g., verbal memory),^19, 20^ sex was also included as a covariate.

**Table 1:**
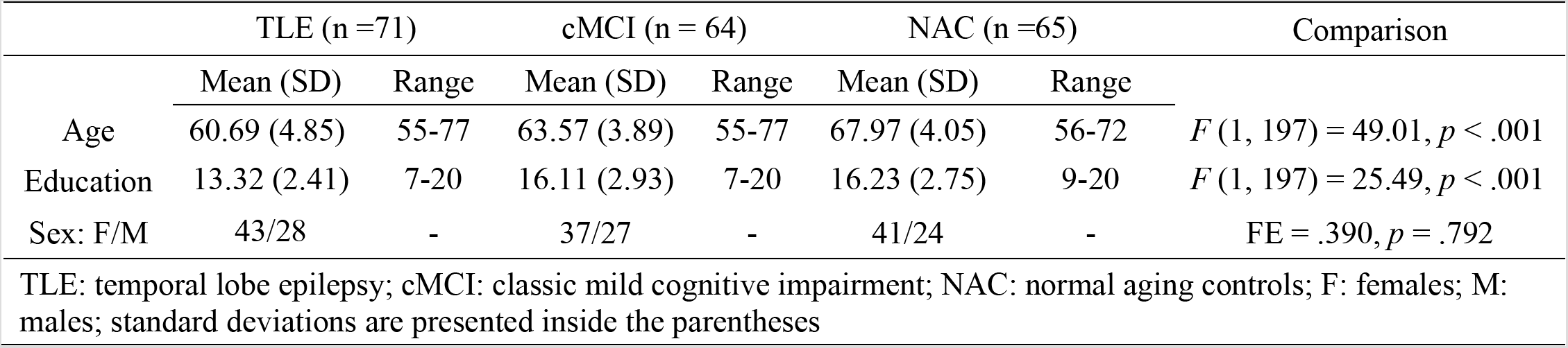
Clinical and demographic characteristics across groups

|  | TLE (n =71) |  | cMCI (n = 64) |  | NAC (n =65) |  | Comparison |
| --- | --- | --- | --- | --- | --- | --- | --- |
|  | Mean (SD) | Range | Mean (SD) | Range | Mean (SD) | Range |  |
| Age | 60.69 (4.85) | 55-77 | 63.57 (3.89) | 55-77 | 67.97 (4.05) | 56-72 | $F(1, 197) = 49.01, p < .001$ |
| Education | 13.32 (2.41) | 7-20 | 16.11 (2.93) | 7-20 | 16.23 (2.75) | 9-20 | $F(1, 197) = 25.49, p < .001$ |
| Sex: F/M | 43/28 | - | 37/27 | - | 41/24 | - | $FE = .390, p = .792$ |

TLE: temporal lobe epilepsy; cMCI: classic mild cognitive impairment; NAC: normal aging controls; F: females; M: males; standard deviations are presented inside the parentheses

### Diagnostic classification in patients with TLE

Forty-three patients (60%) with TLE met criteria for MCI (TLE-MCI), whereas 28 patients did not (40%; TLE-noMCI). Table 2 includes epilepsy-related characteristics/risk factors and CVRFs across TLE-MCI and TLE-noMCI. The only differences between the two patient groups were in age of seizure onset, duration of disease, and history of febrile seizures. Patients in the TLE-MCI group had a younger age of onset, longer duration of disease, and they were more likely to have a history of febrile seizures. When patients were divided into those with an early (< 50 years) versus late (> 50 years) age of seizure onset, 29 patients with an early onset (63%) and 14 patients with a late onset (56%) met criteria for TLE-MCI.

**Table 2:** Clinical and demographic characteristics between TLE-MCI and TLE-noMCI

|  | TLE-MCI | TLE-noMCI | t-value | p-value |
| --- | --- | --- | --- | --- |
| N | 43 | 28 |  |  |
| Age of Onset | 30.02 (20.5) | 42.86 (16.12) | **-2.94\*** | **.005** |
| Duration (years) | 30.58 (20.63) | 18.03 (15.65) | **2.91\*** | **.005** |
| Current # of AEDs | 2.26 (.82) | 1.93 (.86) | 1.62 | .111 |
| # of failed AEDs | 5.67 (2.49) | 5.12 (2.61) | .886 | .379 |
|  |  |  | Fisher's Exact | p-value |
| Sex: Male/Female |  |  |  |  |
| Handedness: L/R/A | 6/35/2 | 1/24/3 | 2.65 | .300 |
| MTS: Y/N | 22/21 | 13/13 | .009 | 1.00 |
| Onset Side: L/R/Bilateral | 25/12/6 | 15/7/5 | .357 | .888 |
| Late Onset+: Y/N | 14/29 | 11/17 | .336 | .617 |
| Monotherapy/Polytherapy | 7/36 | 9/19 | 2.45 | .150 |
| Epilepsy Risk Factors |  |  |  |  |
| Febrile Seizures: Y/N | 7/36 | 0/25 | **8.82** | **.003** |
| Family History of Epilepsy: Y/N | 5/38 | 5/22 | 2.17 | .373 |
| Moderate or Severe TBI: Y/N | 3/40 | 3/24 | 1.94 | .391 |
| Vascular Risk Factors |  |  |  |  |
| Leukoaraiosis: Y/N | 12/29 | 10/15 | .805 | .426 |
| Diabetes: Y/N | 3/40 | 0/28 | 2.04 | .273 |
| Hypertension: Y/N | 14/29 | 10/18 | .075 | .803 |
| Hyperlipidemia: Y/N | 9/34 | 9/19 | 1.13 | .403 |
| BMI > 30: Y/N | 11/28 | 8/18 | .227 | 1.00 |
| Smoking History: Y/N | 14/29 | 12/16 | .775 | .453 |

TLE: temporal lobe epilepsy; MCI: mild cognitive impairment; MI: minimal impaired; F: females; M: males; L: left; R: right; A: ambidextrous; MTS: mesial temporal sclerosis; AEDs: antiepileptic drugs; standard deviations are presented inside the parentheses
\* Equal variance not assumed
+ Late onset: onset of epilepsy 50 or older

The TLE-MCI group was further divided into patients with amnestic (single-domain = 12%; multiple-domain = 60%) and non-amnestic (i.e., language, executive function/processing speed, single-domain = 14%; multiple-domain = 14%) profiles. Figure 1 displays the distribution of T-scores for each measure across TLE-MCI with amnestic profiles, TLE-MCI with nonamnestic profiles, and TLE-noMCI.

### Group differences across cognitive measures

Table 3 includes group comparisons on neuropsychological measures across TLE-MCI, TLE-noMCI, cMCI, and NAC. Figure 2 displays the average performance across the four groups. Overall, TLE-MCI and cMCI were significantly worse on every measure compared to NAC. TLE-MCI and cMCI showed similar performance on TMT-A and TMT-B. TLE-MCI demonstrated worse performance on language measures (BNT, Animal Fluency) relative to cMCI, while the cMCI showed worse performance on memory measures RAVLT-Delayed and RAVLT-Recognition. The TLE-noMCI showed similar performance to NAC on all measures. Compared to TLE-MCI, TLE-noMCI showed similar performance on RAVLT-Recognition, but demonstrated higher scores on all other measures. Relative to cMCI, TLE-noMCI showed similar performance on BNT and Animal Fluency, but higher scores on all other measures.

**Table 3:** Neuropsychological differences across TLE-MCI, TLE-noMCI, cMCI, and NAC.

|  | ANCOVA | TLE-MCI |  |  | TLE- |  |  |
| --- | --- | --- | --- | --- | --- | --- | --- |
|  |  | TLE-MCI<br>vs cMCI | vs TLE-noMCI | TLE-cMCI vs<br>NAC | NoMCI vs<br>cMCI | TLE-noMCI<br>vs NAC | cMCI<br>vs NAC |
| RAVLT-Delayed | **F(3, 170)= 43.52, p<.001** | .006 | **.020** | **<.001** | **<.001** | .358 | **<.001** |
| RAVLT-Recog | **F(3, 166)= 36.29, p<.001** | **.001** | .330 | **.002** | **<.001** | .636 | **<.001** |
| BNT | **F(3, 190)= 10.41, p<.001** | **.025** | .050 | **<.001** | 1.00 | .056 | **.001** |
| Animal Fluency | **F(3, 188)= 23.51, p<.001** | **.022** | **<.001** | **<.001** | .062 | .454 | **<.001** |
| TMT-A | **F(3, 187)= 17.10, p<.001** | 1.00 | **<.001** | **<.001** | **.005** | 1.00 | **<.001** |
| TMT-B | **F(3, 186)= 24.18, p<.001** | 1.00 | **<.001** | **<.001** | **.001** | .899 | **<.001** |

TLE: temporal lobe epilepsy; cMCI: classic mild cognitive impairment; NAC: normal aging controls; RAVLT-Delayed: Rey Auditory Verbal Learning Test delayed; RAVLT-Recog: Rey Auditory Verbal Learning Test recognition; BNT: Boston Naming Test; TMT-A: trail making test condition A; TMT-B: trail making tests condition B Covariates: age, education, sex

**Figure 1:**
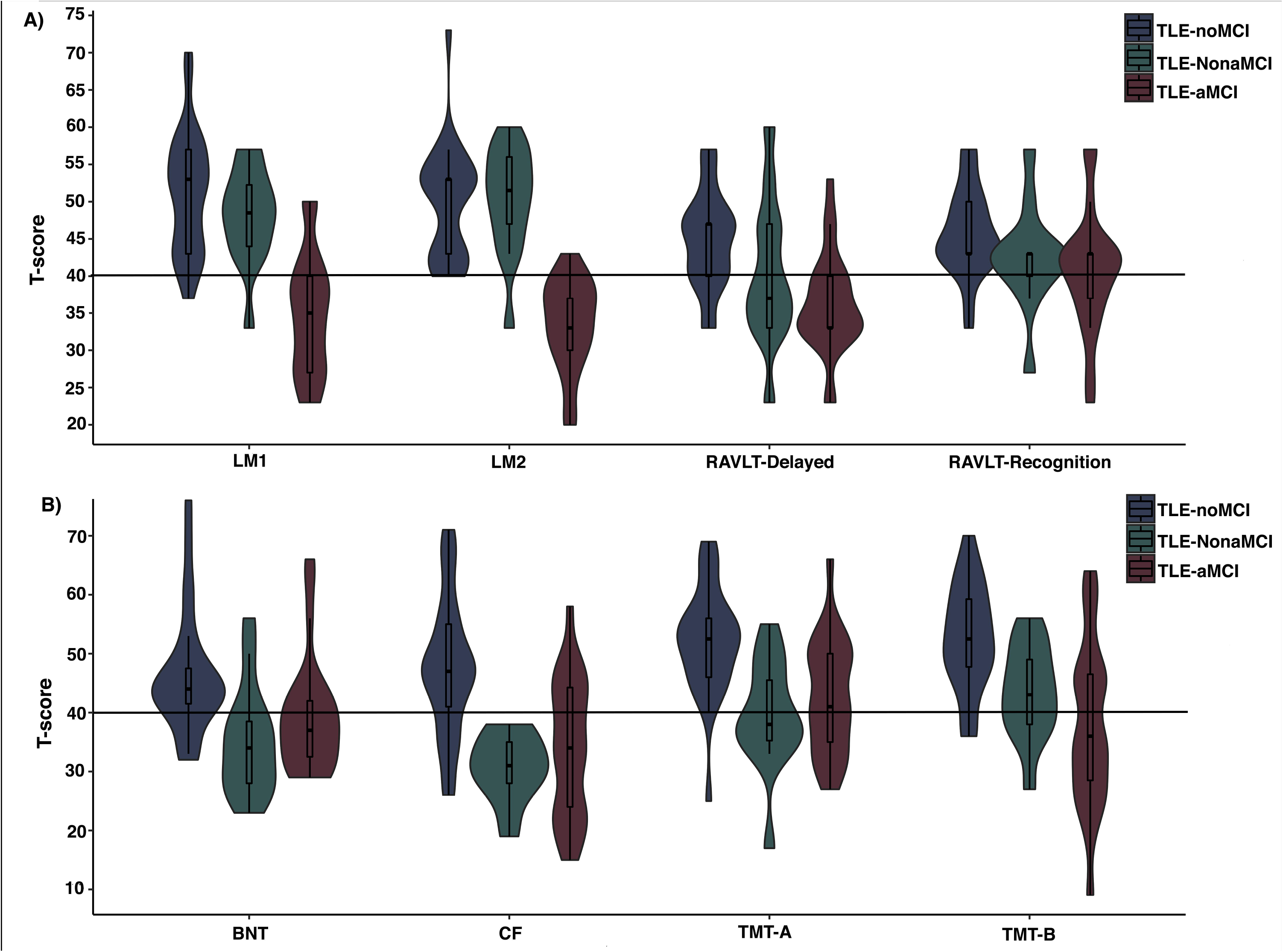
Cognitive scores across TLE subtypes. The distribution of cognitive scores for each neuropsychological test across TLE-MCI with amnestic profiles (TLE-aMCI), TLE-MCI with non-amnestic profiles (TLE-NonaMCI), and TLE-noMCI. The solid line represents impairment at one standard deviation below the mean of a healthy normative sample.

**Figure 2:**
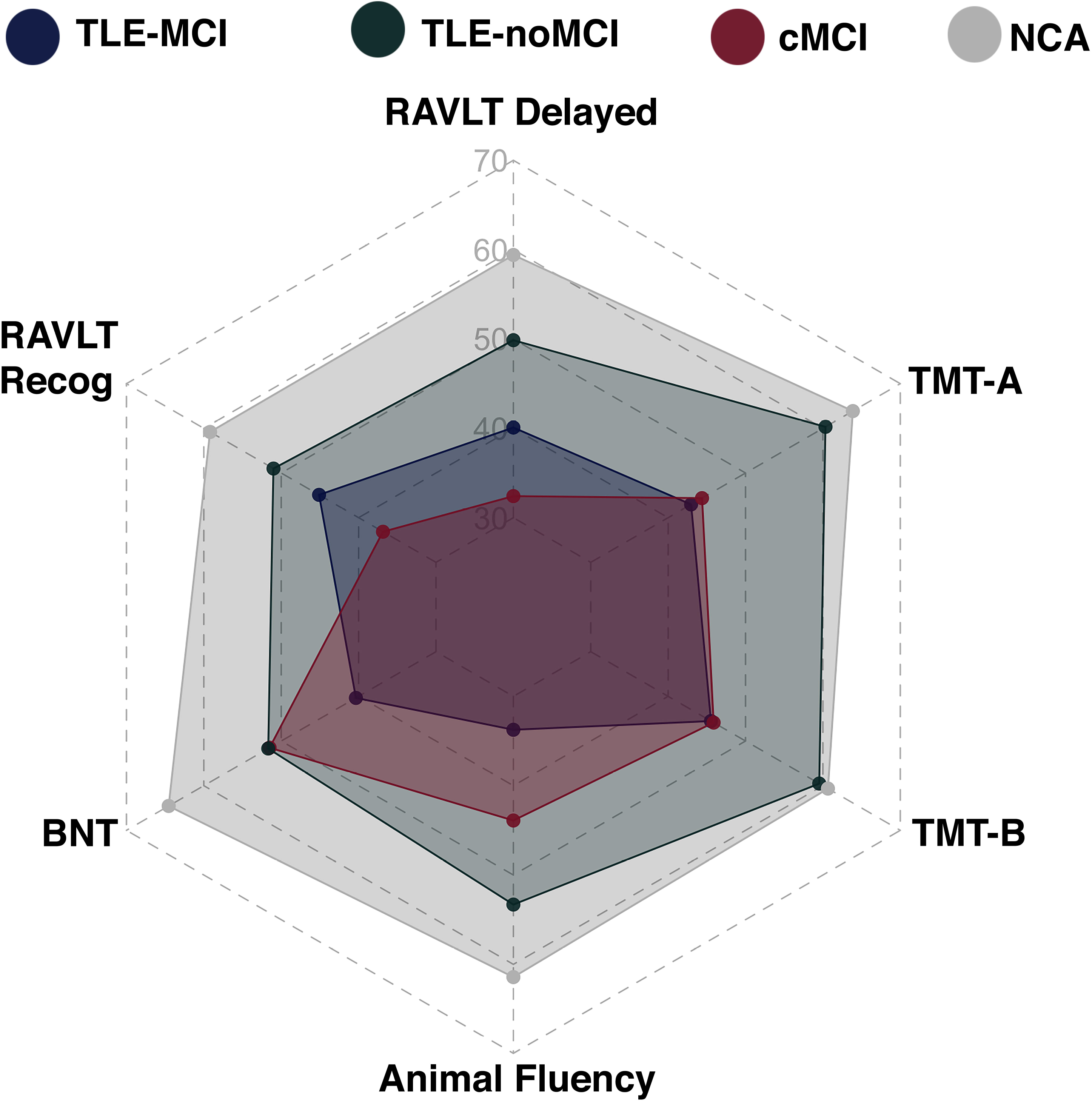
Cognitive profiles across TLE-MCI, TLE-noMCI, cMCI, and NCA. Radar plot demonstrating the overlap in performance across groups. Each point on the plot represents the average performance for each group.

### Contribution of demographic and clinical variables and CVRFs to cognitive impairment in TLE

To reduce the number of variables included in the model, we conducted stepwise regressions to examine the differential contribution of demographics, epilepsy-related clinical variables, and CVRFs to each neuropsychological test for each TLE group. The contribution of age, sex, and education were first evaluated. For TLE-MCI, age was a significant predictor for TMT-A (*β* = −.701, *p* = .040; R^2^ =.115), with increasing age associated with worse processing speed performance. Sex was a significant predictor for RAVLT-Delayed (*β* = 8.98, *p* = .001; R^2^ =.280) and RAVLT-Recognition (*β* = 5.893, *p* = .032; R^2^ =.140), with females performing better than males. Finally, education was a significant predictor for LM1 (*β* = 1.655, *p* = .007; R^2^ =.167), with greater years of education associated with better performance. There were no significant predictors for TLE-noMCI.

The following clinical variables were evaluated for each cognitive test: age of seizure onset, side of seizure onset, MTS status, number of antiepileptic drugs (AEDs), and number of failed AEDs. For TLE-MCI, significant clinical predictors included MTS (*β* = −6.627, *p* = .007; R^2^ = .129) and side of seizure onset (*β* = 4.77, *p* = .008; R^2^change = .165) for RAVLT-Delayed scores, and age of onset (*β* = −.238, *p* = .020; R^2^ = .148) for TMT-B, with left-sided onset, the presence of MTS, and older age of onset associated with worse performance. For TLE-noMCI, the only significant clinical predictor was age of onset for TMT-B (*β* = .270, *p* = .017; R^2^ =.215), with an earlier age of onset associated with worse performance. To evaluate the additional contribution of CVRFs, we controlled for side of onset, MTS status, and age of onset given their contribution to neuropsychological performance described above. This analysis revealed that the presence of leukoaraiosis on MRI was associated with worse performance on RAVLT-Delayed (*β* = −5.841, *p* = .034) in the TLE-MCI group.

### Empirical Classification of TLE-MCI and cMCI

A discriminant function analysis was performed to test whether neuropsychological profiles could correctly classify TLE-MCI and cMCI at the individual subject level. The overall model correctly classified 81.1% of the patients (74.7% with cross-validation) with 90.6% specificity and 61.3% sensitivity (see ROC curve in Figure 3A). Out of the 31 TLE-MCI patients with complete data, 12 patients were mis-classified as cMCI and 6 cMCI patients were misclassified as TLE-MCI (Figure 3B). The distribution of scores across correctly classified and mis-classified patients are presented on Figure 3C. The TLE-MCI mis-classified patients were more likely to have left-sided seizure onset (59%), MTS (N = 7) and leukoaraiosis on MRI (N = 7), and a high prevalence of other CVRFs (N = 10) (see Table 4). As expected, the mis-classified patients demonstrated greater impairments in LM2 (Mean = 33.6), RAVLT-Delayed (Mean = 33.2), and RAVLT-Recog (Mean = 36.4) with more subtle deficits in BNT (Mean = 39.8) and AF (Mean = 37.8).

**Figure 3:**
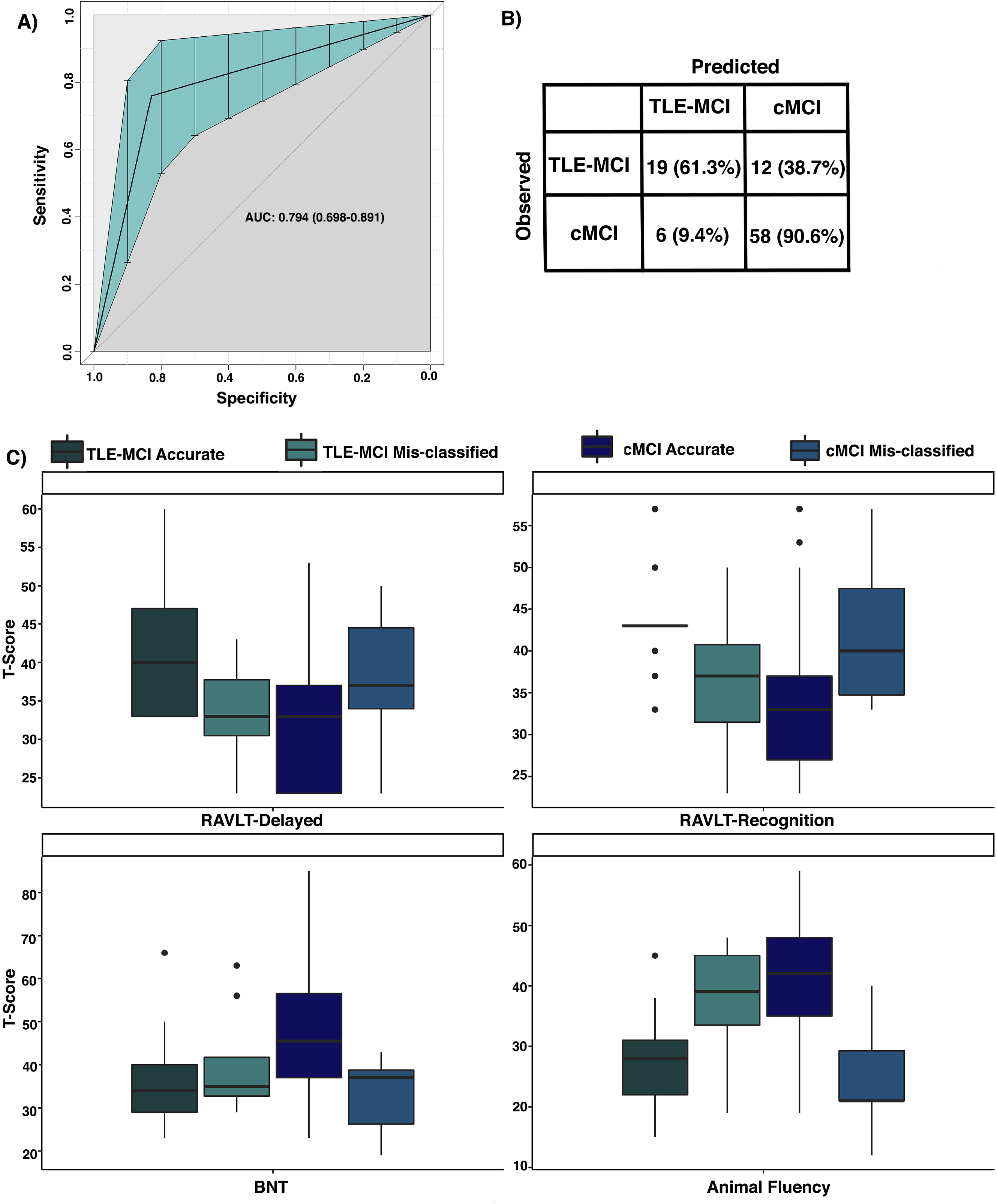
*Empirical Classification of TLE-MCI and cMCI*. A) Receiver operating characteristic (ROC) curve. B) Number of predicted and observed cases for TLE-MCI and cMCI. C) Distribution of scores for patients that were correctly classified and patients that were incorrectly classified.

**Table 4:** Clinical and cognitive characteristics of mis-classified TLE-MCI patients.

| Patient | Onset | Side | MRI | Leukoaraiosis | CVRFs | LM2 | RAVLT-Delayed | RAVLT-Recog | BNT | AF |
| --- | --- | --- | --- | --- | --- | --- | --- | --- | --- | --- |
| 1 | 0 | Left | Other | No | HTN | 53 | 43 | 27 | 56 | 38 |
| 2 | 44 | Right | MTS | Yes | HTN, HLD |  | 23 | 43 | 35 | 32 |
| 3 | 52 | Left | MTS | No | HLD | 20 | 23 | 27 | 37 | 19 |
| 4 | 5 | Left | MTS | No | HTN, HLD | 33 | 37 | 37 | 32 | 34 |
| 5 | 53 | Right | Nonlesional | No | DM, Obesity | 37 | 33 | 23 | 63 | 45 |
| 6 | 10 | Left | Other | No | - | 20 | 33 | 33 | 35 | 41 |
| 7 | 56 | Left | Encephalomalacia | Yes | DM | 40 | 33 | 50 | 29 | 45 |
| 8 | 11 | Bilateral | MTS | Yes | - | 30 | 40 | 37 | 33 | 40 |
| 9 | 28 | Right | MTS | Yes | HTN, Obesity | 37 | 37 | 40 | 56 | 47 |
| 10 | 44 | Left | MTS | Yes | HLD | 27 | 23 | 40 | 30 | 34 |
| 11 | 26 | Bilateral | MTS | Yes | HTN, Obesity | 33 | 40 | 43 | 34 | 48 |
| 12 | 55 | Left | Other | Yes | HTN, HLD, Obesity | 40 | 33 | 37 | 37 | 31 |

MTS: mesial temporal sclerosis; HTN: hypertension; HLD: hyperlipidemia; DM: diabetes mellitus; LM2: Logical Memory 2; RAVLT: Rey Auditory Verbal Learning Test; Recog: recognition; BNT: Boston Naming Test; AF: Animal fluency

## Discussion

Despite older adults with TLE representing a rapidly growing cohort of patients with epilepsy, the neuropsychological profile of these patients has not been comprehensively characterized. Furthermore, there is no consensus on diagnostic criteria for cognitive disorders associated with aging in TLE, or epilepsy in general. This is concerning given the large number of patients meeting criteria for TLE-MCI in our study, a sizable number of who may be at increased risk for dementia^3^. A uniform definition will help researchers and clinicians, 1) stratify the risk for dementia, 2) study the effects of cognitive impairments on quality of life and functional independence, 3) improve long-term health outcomes associated with aging, and 4) converse in a standard language regarding cognitive diagnoses using clear opertational criteria.

We provide the first characterization of the nature and prevalence of a cognitive disorder in a large and well-characterized group of older adults with TLE. We demonstrate that approximately 60% of older TLE patients meet diagnostic criteria for a cognitive disorder. Second, we demonstrate that these patients exhibit neuropsychological profiles that are similar, but not identical, to patients with cMCI. In fact, 61% of patients with TLE-MCI could be distinguished from those with cMCI based on their language and memory profiles. Finally, we demonstrate that both epilepsy-related factors and CVRFs may play an important role in their impairment profiles. These data suggest that over half of older adults with TLE meet clinical criteria for MCI, but that the nature of this impairment in a majority of patients is phenotypically different from prodromal AD and may reflect underlying epileptogenic and vascular pathology, along with changes secondary to years of on-going seizures.

## The TLE-MCI Phenotype

Decades of research in epilepsy has demonstrated that cognitive dysfunction is a highly prevalent and debilitating comorbidity in TLE^21, 22^. However, the TLE literature has mainly focused on characterizing the cognitive trajectories of children and young to middle-aged adults and very little is known about cognitive impairment in older adults^2^. Critically, the prevalence of clinical diagnoses associated with abnormal cognitive aging (e.g., MCI, dementia) remains unknown. A small number of empirical studies have compared the average cognitive profiles of older patients with epilepsy to healthy controls and have reported worse cognitive performance in epilepsy patients^8, 9, 23–25^. However, these studies have included heterogeneous groups of patients with a wide range of epilepsy syndromes and have not applied uniform diagnostic criteria. Here, we demonstrate that older adults with TLE who meet diagnostic criteria for MCI (TLE-MCI) harbor significant cognitive impairment characterized by an amnestic, multi-domain profile with deficits most commonly in memory and language. By applying widely used empirically-derived diagnostic criteria for MCI, we were able to identify the presence of a cognitive disorder in 63% of patients with long-standing TLE and 56% of those with a more recent epilepsy diagnosis. There are studies suggesting progressive cognitive decline in patients with long-standing TLE^21^, as well as studies documenting the presence of cognitive deficits around the time of diagnosis in patients with late onset epilepsy that eventually follow a progressive course^8, 26^. Thus, a diagnostic approach such as the one employed in our study offers the ability to identify patients that may be at risk for further cognitive decline and/or dementia and may benefit from early intervention (e.g., control of CVRFs) and close monitoring of their cognitive functioning.

Notably, when comparing the cognitive profiles of TLE-MCI to cMCI, an interesting pattern emerged. The TLE-MCI were highly similar to those with cMCI on measures of verbal learning, processing speed and executive functioning. Unique to TLE-MCI were impairments in language, whereas cMCI demonstrated evidence of more rapid forgetting. Importantly, we were able to correctly classify 81% of the patients based on cognitive scores alone. Therefore, a majority of TLE-MCI patients appear phenotypically different from cMCI, which could have diagnostic value and implications for differentiating their cognitive trajectories and risk for progression to AD.

## Similarities in the TLE-MCI phenotype and cMCI

In addition to the shared cognitive dysfunction observed in cMCI and older adults with TLE^9^, there is evidence suggesting a bi-directional relationship between TLE and AD. For instance, patients with epilepsy are at increased risk for developing AD^27^, and patients with AD have a six- to tenfold higher risk for developing seizures^28^. This bi-directional relationship has been linked to the presence of tau pathology^29^, amyloid-B precursor protein^30^, and senile plaques^31, 32^ in TLE, all of which are pathological hallmarks of AD. Nardi Cesarini et al.^8^, reported that patients with late-onset epilepsy of unknown origin who met criteria for MCI, had similar AD-like cerebrospinal fluid profiles to patients with MCI without seizures. However, AD-related biomarkers have also been identified in resected tissue of young to middle-aged TLE patients with *chronic* epilepsy. Therefore, it is unclear whether deposition of AD-related pathology contributes to the development of seizures, is the consequence of many years of seizures^3^, or is unrelated to epilepsy. Although we were able to correctly classify 81% of the patients based on cognitive scores alone, approximately 39% of TLE-MCI patients were misclassified as cMCI. This is of interest given that a subset of older adults with TLE and AD-related pathology and progress to AD. Interestingly, these mis-classified patients include both those with early and late seizure onsets as well as elevated CVRFs. Thus, it is possible that this subset of patients with a more classically MCI-like cognitive profile may represent a sub-group that is on a progressive course to AD. There are no established recommendations for biomarkers or neuropsychological tests specific for older adults who present first with epilepsy but may harbor a progressive neurodegenerative disorder. Therefore, MCI may be missed in these patients as cognitive impairments are attributed to their known seizure disorder and comorbid disorders are often overlooked.

## Contribution of epilepsy and CVRFs to cognitive impairment

We found that the presence of MTS and left-sided seizure onset was associated with worse cognitive performance in delayed memory and executive function. These clinical variables have been shown to impact cognition and predict long-term cognitive and post-operative outcomes^6^. In addition, we found that female sex, greater years of education, and younger age were associated with better performance across different cognitive measures. Taken together, we demonstrate that in TLE-MCI patients there are important clinical and demographic features that contribute to their cognitive profiles. It is possible that with advancing age these clinical features further exacerbate an already diminished cognitive capacity and could be used to identify the patients at increased risk for further cognitive decline.

In our cohort, approximately 60% of TLE patients had at least one CVRF and 31% of patients had evidence of leukoaraiosis on MRI. After controlling for important epilepsy-related clinical variables, leukoaraiosis was associated with poorer delayed memory performance. This pattern emerged in patients with both early and late onset seizures, suggesting that vascular pathology may lead to a worsening of pre-existing memory impairments in older adults with epilepsy. A 50-year follow up population-based study, revealed increased MRI markers of cerebrovascular disease in adults with childhood-onset epilepsy^33^. Thus, patients with early-onset epilepsy may start with a greater cerebrovascular risk when they reach middle-age and those with a late-onset may already have a diminished brain and cognitive reserve that have resulted in the clinical manifestation of seizures. However, the cumulative effect of CVRFs and epilepsy-related factors is not well understood. Prospective longitudinal studies are needed to identify modifiable risk factors that could mitigate cognitive and functional decline, and potentially halt progression to dementia in high risk patients.

## Limitations

Although the tests included in our study are among the most commonly used measures in epilepsy clinics, we did not include measures designed to assess for cognitive decline in older adults that are commonly used in memory-disorders clinics. However, our test selection is representative of common clinical practice and can be used to identify patients who may benefit from referral to a memory disorders/dementia clinic. As the number of older adults with epilepsy continues to increase, neurologists and neuropsychologists who see patients with epilepsy will need updated guidelines on the diagnostic and treatment of older adults with epilepsy who may also be presenting with cognitive deficits suggestive of a progressive disorder of aging. Second, our study is cross-sectional, and we did not have longitudinal data or information on progression to dementia. Longitudinal studies of older patients with early and late onset epilepsy are greatly needed in order to risk-stratify patients for progression to dementia and implement early interventions aimed at delaying the negative impact of cognitive decline on quality of life and functional independence. Finally, we did not have biomarker or genetic data on our epilepsy cohort given that they are not routinely collected as part of standard medical care. Collecting biomarkers and genetic data could help to further identify patients in the prodromal state of AD.

## Conclusion

We demonstrate that 60% of patients with TLE who are over the age of 55 meet diagnostic criteria for a cognitive disorder when comprehensive neuropsychological criteria are applied. As the field of epilepsy moves toward *precision neuropsychology*, it is critical to develop a diagnostic framework that can be applied to older adults with epilepsy across clinics and geographical locations. The diagnostic method applied in our study in the first step in this direction, providing an operational definition to impairment that can be used with different neuropsychological batteries.

## Data Availability

Authors have full access to all study data and participant
consent forms and take full responsibility for the data, the
conduct of the research, the analysis and interpretation of the
data, and the right to publish all data.

## Acknowledgements

Supported by NIH/NINDS R01 NS065838 (CRM); T32 MH018399 (EK); F31 NS111883-01 (AR); R01 NS088748 and K02NS070960 (DLD); R01NS111022 and R01 AG027161-11 (BPH); NIH/NINDS R01NS097719 and R01NS035140 (RMB); and the U.S. Department of Veterans Affairs CSR&D Service (CDA-2 1IK2CX001415; ECE). Dr. Herman receives support by grants from Citizens United for Research in Epilepsy. Dr. Drane receives grant/contract funding from Medtronic, Inc.

## Author Contributions

Conception and design of the study: A.R., and C.R.M. Acquisition and analysis of data: A.R., D.L.D., V.P., R.M.B., B.P.H., and C.R.M. Drafting a significant portion of the manuscript or figures: A.R., E.K., E.C.E., A.C.M., Z.I.W., D.L.D., V.P., R.M.B., B.P.H., and C.R.M.

## ADNI

Data collection and sharing for this project was funded by the Alzheimer’s Disease Neuroimaging Initiative (ADNI) (National Institutes of Health Grant U01 AG024904) and DOD ADNI (Department of Defense award number W81XWH-12-2-0012). ADNI is funded by the National Institute on Aging, the National Institute of Biomedical Imaging and Bioengineering, and through generous contributions from the following: AbbVie, Alzheimer’s Association; Alzheimer’s Drug Discovery Foundation; Araclon Biotech; BioClinica, Inc.; Biogen; Bristol-Myers Squibb Company; CereSpir, Inc.; Cogstate; Eisai Inc.; Elan Pharmaceuticals, Inc.; Eli Lilly and Company; EuroImmun; F. Hoffmann-La Roche Ltd and its affiliated company Genentech, Inc.; Fujirebio; GE Healthcare; IXICO Ltd.; Janssen Alzheimer Immunotherapy Research & Development, LLC.; Johnson & Johnson Pharmaceutical Research & Development LLC.; Lumosity; Lundbeck; Merck & Co., Inc.; Meso Scale Diagnostics, LLC.; NeuroRx Research; Neurotrack Technologies; Novartis Pharmaceuticals Corporation; Pfizer Inc.; Piramal Imaging; Servier; Takeda Pharmaceutical Company; and Transition Therapeutics. The Canadian Institutes of Health Research is providing funds to support ADNI clinical sites in Canada. Private sector contributions are facilitated by the Foundation for the National Institutes of Health (https://www.fnih.org). The grantee organization is the Northern California Institute for Research and Education, and the study is coordinated by the Alzheimer’s Therapeutic Research Institute at the University of Southern California. ADNI data are disseminated by the Laboratory for Neuro Imaging at the University of Southern California.

